# Interactive effects of genetic variants and oral contraceptive use on depression in the UK Biobank

**DOI:** 10.64898/2026.08.05.26359775

**Authors:** Clair A. Enthoven, Rosa H. Mulder, Alexander Neumann, Therese Johansson, Frances S. Chen

## Abstract

**Background:** Oral contraceptive (OC) use, particularly during adolescence, may increase depression risk in some individuals, but it remains unclear who is susceptible to mood-related side effects and who is not. We aimed to detect single nucleotide polymorphisms (SNPs) and genes that moderate the effect of OC use on depression in young adulthood using data from the UK Biobank.

**Methods:** N=202,243 participants were followed from birth to age 23.29 (SD: 2.58) years. We used Cox models for counting processes to test the association between OC use and incident depression in young adulthood, and conducted a genome-wide-by-drug-interaction study (GWDIS) of SNP by OC use interactions alongside a standard genome-wide association study (GWAS) on incident depression in young adulthood.

**Results:** Over half of all participants (57.5%) initiated OC and 1.0% received a depression diagnosis during follow up. OC initiators had a 20% higher hazard of incident depression than non-initiators (HR=1.20, 95% CI=1.04-1.37). No SNPs reached genome-wide significance in the GWDIS, though eight showed suggestive interaction signals (p<1e-5). At the gene level, FSIP1 (p=5.90e-5) and EHBP1 (p=6.47e-5) showed suggestive signals, but none passed the genome-wide threshold. No SNPs reached genome-wide significance in the GWAS.

**Conclusions:** We did not find evidence for genetic variants that moderate the association between OC initiation and depression. If such effects exist, they are likely to be small and polygenic, suggesting there is currently no solid basis for using genetic data for individualised contraception counselling concerning mood-based side effects.

## Introduction

Depressive disorders are highly prevalent worldwide, affecting more than 300 million people, with global rates continuing to rise.^1,2^ Onset commonly occurs during adolescence and early adulthood, and adult women are affected at approximately twice the rate of men.^3^ Although the reasons for these pronounced sex differences are not yet fully understood, they are likely driven by a complex interplay of environmental and biological factors.^4^ Recent genetic research underscores this complexity: the largest genome-wide association study (GWAS) of depression to date identified 697 genetic variants and 308 associated genes, while sex-stratified GWAS analyses suggest that distinct genetic pathways may underlie depression risk in males and females.^5,6^

Women’s heightened vulnerability to anxiety and depression is closely linked to reproductive transitions (e.g., puberty, the peripartum period, and perimenopause), implicating a role for sex hormones including estrogens and progesterone.^7^ Hundreds of millions of people worldwide use oral contraceptives (OC), which typically contain synthetic forms of estrogens and progesterone.^8^ While the majority of users experience few or no adverse effects, approximately 4–10% of the users report negative mood-related side effects, including depression, anxiety, and irritability.^9^ These effects can be severe and represent one of the most common reasons for switching hormonal contraceptive formulations or discontinuing use altogether.^9^

Recent research has also begun to deliver more direct evidence that the use of hormonal contraceptives, especially during adolescence, may heighten risk for depression and suicide.^10–13^ For example, nationwide cohorts link hormonal contraceptive use to first-time antidepressant use and depression diagnosis,^13^ as well as to suicide attempt and completed suicide,^10^ and prospective and retrospective cohort studies report similar adolescent-specific signals for depression in (early) adulthood.^11,12^ However, other studies, including a meta-analysis of randomized trials, prospective and cross-sectional cohort studies have found no relationship.^14–16^ Finally, some prospective cohort studies reported a negative association, indicating less depressive symptoms among adolescent hormonal contraceptive users as compared to non-users.^17–19^

The sources of the discrepancies in existing findings are multifaceted. For example, some studies compare OC users versus non-users without taking history of OC use into account, which may lead to a healthy user bias.^20,21^ In addition, women who tolerate OC well are more likely to participants in randomized trials.^21^ Moreover, a lack of randomization in observational studies may lead to residual confounding when studies fail to adjust for important confounders. However, the discrepancies in findings also highlights the likelihood that hormonal contraception may affect mental health in some individuals, but not in others,^22^ which is underscored by the high heterogeneity shown in meta-analyses.^14,23–26^ Since the onset of mental disorders and the initiation of hormonal contraception often coincide during adolescence and young adulthood, causal interpretations are challenging. Using genetic data in predicting which individuals may be more vulnerable to experiencing mental health changes following the use of hormonal contraception could enhance personalised approaches to contraception counselling.

There are several plausible mechanisms through which genetic factors could moderate OC effects on depression. Firstly, people differ in the rate at which they metabolize sex hormones,^27^ likely in part due to genetic variability affecting the availability of liver enzymes that are encoded by the cytochrome P450 gene family (*CYP*).^28^ Secondly, genetic variability of sex hormone receptors such as *ESR1* and *ESR2* has been linked to depression and social functioning.^29,30^ Finally, research on the mineralocorticoid receptor gene (*MR*) has shown that female carriers of MR-haplotype 2 have a lower risk of depression during their reproductive years,^29^ and they were less sensitive to mood-effects of OC.^30^

Thus far, most studies have restricted their focus to a few candidate genes only, due to limited power for genome-wide approaches. While these smaller, hypothesis-driven studies can enhance our understanding of specific gene-drug interaction effects, they frequently fall short in uncovering novel disease-causing mechanisms and tend to produce findings that are difficult to replicate.^31^ In this study, we conducted a genome-wide-by-drug-interaction study (GWDIS) as well as a genome-wide-association study (GWAS) to detect genetic markers linked to depression in young adulthood. Specifically, we examined whether genetic variants moderate the effect of OC use on depression diagnosis before the age of 25 years.

## Methods

### Study population

UK Biobank is a prospective biomedical database containing genetic, lifestyle, and health information of over 500,000 individuals in the UK.^32^ Recruitment began in 2006, and new data are uploaded into the database regularly. N=501,939 participants provided informed consent. We excluded participants who did not have whole genome sequencing data available (N=17,060), participants with male sex (N=221,957; UKB field: 31), and participants with a sex mismatch (N=82; UKB field: 22001). To minimize confounding due to population stratification, we restricted the sample to participants who self-identified as ‘White British’ and have very similar genetic ancestry based on a principal components analysis of the genotypes (N=42,950 were excluded; UKB field: 22006). Participants were excluded when their genetic data had outliers for heterozygosity (N=370; UKB field: 22027). Furthermore, we excluded participants with missing data on OC use and age at first OC use (N=6,732). Out of all participants with shared relatedness of up to the third degree (UKB field: 22021; kinship coefficients >0.044), only one participant was included using the ukb_gen_samples_to_remove function of the ukbtools R package (N=10,371 were excluded).^33^ We also excluded N=165 participants who initiated OC in the same year they were diagnosed with depression, because the order of occurrences could not be determined. Finally, we excluded N=1 participant who reported a depression at age 0, leaving an analytical sample of N=202,243 participants. The construction of the analytical sample is shown in Supplementary Figure 1. The UKB study was approved by the North West - Haydock Research Ethics Committee (16/NW/0274) and this project was preregistered at Open Science Framework (https://doi.org/10.17605/OSF.IO/ZE3KY). Deviations from the pre-registration are described in Supplementary Table 1.

**Figure 1.**
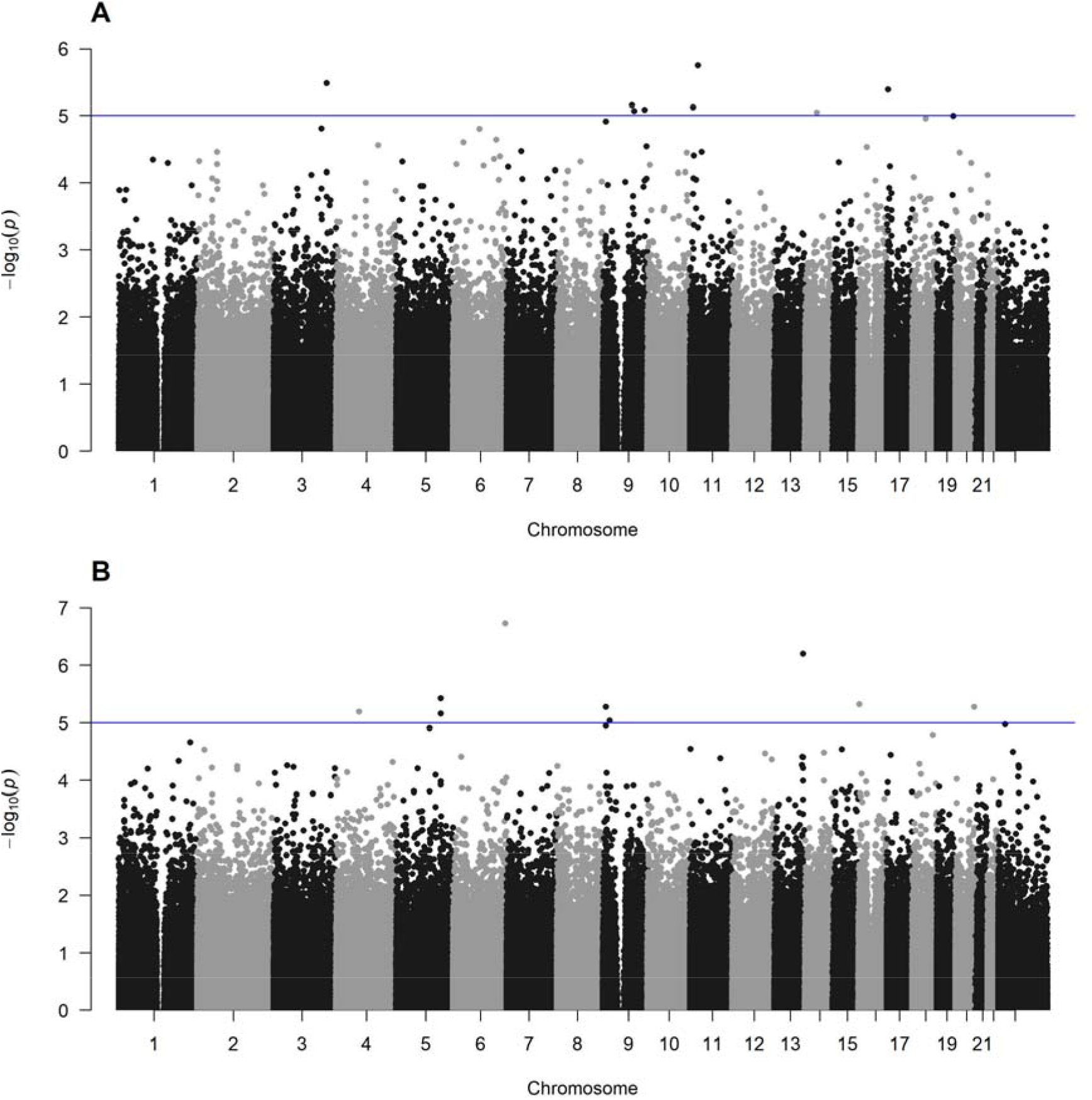
A and B: Manhattan plot of genome-wide interaction (A) and genome-wide association (B) p-values. The blue horizontal line indicates p-value < 1e-5.

### OC use

Information on OC use (UKB field: 2784) and age at initiation of OC use (UKB field: 2794) was obtained through a questionnaire during the initial assessment visit. The majority of the participants initiated OCs in 1970s and early 1980s when the second-generation pills containing ethynyl-estradiol and levonorgestrel were predominantly used in the United Kingdom.^34^ Age at OC initiation was processed as a time dependent variable.

### Incident depression

Information on the first diagnosis of depression was obtained from either the verbal interview during the initial UK Biobank assessment visit or the International Classification of Disease code F32 recorded in the inpatient hospital or primary care data. During the initial UK Biobank assessment visit, participants were asked whether they had ever been told by a doctor that they had an illness or disability. The interviewer showed a tree structure of diagnoses which was loosely based on the ICD-10 codes. For each illness the participant recorded, the interviewer recorded their age or the year of the first diagnosis. If a participant had both an ICD-10 code identified in the registers (i.e., data from either the death register, hospital inpatient admissions, or primary care) and a self-reported medical condition, the earliest date at which a depression diagnosis was identified in any of these sources was used (UKB field: 130894).^6,34,35^ Age at first depression was calculated by subtracting the participant’s year of birth (UKB field: 34) from the year of first depression diagnosis.

### Covariates

Covariates in the UK Biobank were selected based on literature and consisted of year of birth (UKB field: 34),^36^ Townsend deprivation index (UKB field: 22189),^17,37^ age at menarche (UKB field: 2714),^38,39^ age at sexual debut (UKB field: 2139),^17,40^ age at first reported acne (UKB field: 131790), ovarian dysfunction (UKB field: 130736),^41,42^ endometriosis (UKB field: 132122),^43,44^ dysmenorrhea (UKB field: 132146),^45,46^ the first eight genetic principal components (UKB field: 22009),^6^ and genotyping array (UKB field: 22000). More information on the covariates is provided in Supplementary Table 1.

### Genotyping, imputation and quality control

DNA was extracted from stored blood samples collected from participants during their study visit. Detailed array design, genotyping, and quality control procedures have been described previously.^32^ In brief, the phasing and imputing were performed using the Haplotype Reference Consortium (HRC) and additionally with the merged UK10K and 1000 Genomes phase 3 reference panels. The imputed data was then combined, using HRC imputation when a SNP was present in both panels.^32^ Whole Genome Sequencing data for 500,000 participants was re-processed using DRAGEN 3.7.8, with individual and joint-called data available for all participants. Quality control was performed with PLINK v2.0.^47^ We only included variants with the PASS filter indicating a site quality score (MLSQ) ≥0.1 and a genotype rate ≥90% recommended by the data providers.^48^ Furthermore, we removed variants with a minor allele frequency <5%, minor allele count <25, Hardy-Weinberg equilibrium test P<1e-100 and we only included SNPs, leaving a total of 6,200,815 SNPs. Finally, we used linkage disequilibrium (LD)-based marker pruning with a window size of 200 kilobase and an r^2^ threshold of 0.5 leaving a total of 518,643 SNPs for analyses.

### Statistical analyses

Prior to analyses, single imputations were performed to impute missing data on the covariates using the principal component method “factorial analysis for mixed data” of the missMDA R package.^49^ The percentage of missing data ranged from 0% to 10.5% (Table 1). Participants were followed from birth until age 25, the first occurrence of depression, their first pregnancy (UKB fields age at birth minus one: 2754 and 3872), if they reached menopause (UKB field: 3581), underwent a hysterectomy (UKB field: 2824) or a bilateral oophorectomy (UKB field: 3882) whichever came first. OC initiation was processed as a time dependent variable using Cox modelling for counting processes.^50^ Firstly, we tested the association of OC use on incident depression in adolescence and young adulthood.^34^ Secondly, we conducted a genome-wide association study (GWAS) on incident depression in adolescence and young adulthood. Thirdly, we conducted a genome-wide-by-drug-interaction study (GWDIS) with OC use on incident depression in adolescence and young adulthood. All analyses included OC initiation and were adjusted for all covariates. Age at menarche, sexual debut and first reported acne were processed as time dependent covariates, while the others were processes as fixed covariates. The GWDIS included a SNP x OC use interaction term, and the analyses were conducted in R.

**Table 1:** General characteristics of the cohort.

|  | **Non-initiators**<br>**(N=85,908)** | **OC initiators**<br>**(N=116,335)** | **Total**<br>**(N=202,243)** | **p-value** |
| --- | --- | --- | --- | --- |
| **Depression; N (%)** | 1,341 (1.6%) | 726 (0.6%) | 2,067 (1.0%) | < 0.001 |
| N missing data | 0 | 0 | 0 |  |
| **Source depression diagnosis** |  |  |  | < 0.001 |
| Primary care and other source(s) | 34 (2.5%) | 28 (3.9%) | 62 (3.0%) |  |
| Primary care only | 78 (5.8%) | 92 (12.7%) | 170 (8.2%) |  |
| Self-report and other source(s) | 550 (41.0%) | 255 (35.1%) | 805 (38.9%) |  |
| Self-report only | 679 (50.6%) | 351 (48.3%) | 1,030 (49.8%) |  |
| **Age menarche; Mean (SD)** | 12.96 (1.62) | 12.95 (1.59) | 12.95 (1.60) | 0.444 |
| N no menarche/after follow-up | 170 | 2 | 172 |  |
| N missing data | 2,098 | 2,356 | 4,454 |  |
| **Age sexual debut; Mean (SD)** | 18.93 (2.69) | 18.34 (2.49) | 18.55 (2.58) | < 0.001 |
| N no sexual debut/after follow-up | 10,932 | 966 | 11,898 |  |
| N missing data | 13,792 | 7,403 | 21,195 |  |
| **Age first acne; Mean (SD)** | 18.08 (3.60) | 17.97 (3.87) | 18.00 (3.79) | 0.698 |
| N no acne/after follow-up | 85,649 | 115,762 | 201,411 |  |
| N missing data | 0 | 0 | 0 |  |
| **Year of Birth; Mean (SD)** | 1946.96 (6.77) | 1954.66 (6.98) | 1951.39 (7.87) | < 0.001 |
| N missing data | 0 | 0 | 0 |  |
| **Neighborhood deprivation; Mean (SD)** | -1.38 (3.00) | -1.78 (2.76) | -1.61 (2.87) | < 0.001 |
| N missing data | 76 | 158 | 234 |  |
| **Ovarian dysfunction; N (%)** | 507 (0.6%) | 1,143 (1.0%) | 1,650 (0.8%) | < 0.001 |
| N missing data | 0 | 0 | 0 |  |
| **Endometriosis; N (%)** | 2,540 (3.0%) | 5,063 (4.4%) | 7,603 (3.8%) | < 0.001 |
| N missing data | 0 | 0 | 0 |  |
| **Dysmenorrhea; N (%)** | 6,706 (7.8%) | 15,305 (13.2%) | 22,011 (10.9%) | < 0.001 |
| N missing data | 0 | 0 | 0 |  |
| **Follow-up time; Mean (SD)** | 22.11 (3.09) | 24.16 (1.67) | 23.29 (2.58) | < 0.001 |
| N missing data | 0 | 0 | 0 |  |

*N=number of participants; SD=standard deviation.*

The gene-by-OC-interaction (GxOC) variance component and the oracle polygenic score-by-OC-interaction (PGSxOC) were estimated using polygenic gene-environment interaction linkage disequilibrium score regression (PIGEON-LDSC) from https://github.com/qlu-lab/PIGEON.^51^ The GxOC variance component indicates the proportion of phenotypic variance explained by gene-OC interaction, while the oracle PGSxOC between OC use and a polygenic depression score based on the GWAS (without SNP x OC use interaction term) estimates the covariance between the effects of genetic predisposition and OC use, therefore indicating the direction and degree of the interaction on genome-wide scale.^51^ Finally, the autosomal SNP-based heritability (h^2^_SNP_) of the GWAS was estimated using linkage disequilibrium score regression (LDSC) from https://github.com/CBIIT/ldsc.^52^ For all analyses, the linkage disequilibrium structure was estimated using the 1000G EUR reference population from https://doi.org/10.5281/zenodo.7768713, and a population prevalence of 0.1 was used to calculate the liability scales.

We conducted Functional Mapping and Annotation of genes using the FUMA SNP2GENE function to identify lead/independent SNPs and annotated genes.^53^ The summary statistics of the GWAS and GWDIS were used as input, and were lifted from GRCh38 to GRCh37 using liftOver() function from the rtracklayer R package based on the UCSC hg38ToHg19 chain file.^54,55^ Variants without unique mappings (n=1,997) or with mapping on the Y chromosome (n=1) were excluded. Since none of the SNPs reached genome-wide significance (p-value < 5e-8), the maximum *P* value for lead SNPs was set to 1e-5, and the r^2^ threshold for independent SNPs was set to 0.6. The UK Biobank release 2b 10k White British was used as reference panel population. Positional mapping was done for SNPs located within a 10 kb window upstream or downstream of a gene. Finally, we conducted a gene-level analysis using MAGMA.^53,56^

## Results

### General characteristics

Participants (N=202,243) were born between 1936 and 1970 with mean birth year 1951.39 (SD: 7.87). The mean follow-up time from birth to the first occurrence of depression, first pregnancy, menopause, hysterectomy or bilateral oophorectomy or age 25 years was 23.29 (SD: 2.58) years. During the follow-up time, N=116,335 (57.5%) initiated OC at mean age 19.55 (SD: 2.60) years, and a total of N=2,067 participants (1.0%) had a depression diagnosis at mean age 18.88 (SD: 4.00) years. Participants who initiated OC during the follow-up time were younger, had an earlier sexual debut, lived in a less deprived neighborhood, had more often ovarian dysfunction, endometriosis and/or dysmenorrhea and they had a longer follow-up time as compared to non-users (Table 1). Participants who initiated OC had an increased risk of a later depression diagnosis compared to participants who did not initiate OC in both the unadjusted analyses (HR_unadjusted_=1.66; 95% CI=1.48-1.86) and the adjusted analyses (HR_adjusted_=1.20; 95% CI=1.04-1.37).

### Genome-wide-by-drug-interaction study findings

None of the SNPs from the GWDIS reached genome-wide significance (Figure 1A). Table 2 summarizes the eight SNPs with a p-value < 1e-5 that were carried forward for Functional Mapping and Annotation analyses. Six of them were independent lead SNPs, and four of them were mapped to several genes. None of the SNPs from the GWDIS had relatively small p-values in the GWAS (p>0.44), and vice versa (p>0.35). Figure 2A shows the QQ plots of observed versus expected p-values, which closely followed the null distribution (λ=1.00).). The oracle PGSxOC estimate indicates that OC use effects are stronger with higher genetic predisposition for depression, however, the interaction was small and not statistically significant (α_i_= 0.08, SE = 0.12, p = 0.48). This observation is further strengthened by a negative proportion of phenotypic variance explained by gene-OC interaction (GxOC variance component on the liability scale was −0.06 (SE = 0.07, p = 0.41).

**Table 2:** SNPS with a p-value < 1e-5 in the genome-wide-by-drug-interaction study on depression in young adulthood. None of the SNPs passed the genome-wide significance threshold (< 5e-8).

| Marker | rsID | MAF | A1/A2 | Effect<br>t<br>GWDI<br>S | p-value<br>GWDIS | Effect<br>t<br>GWAS | p-value<br>GWAS | Independent<br>SNP | Mapped genes |
| --- | --- | --- | --- | --- | --- | --- | --- | --- | --- |
| 11:25992172 | rs117355255 | 0.069 | A/G | -0.706 | 1.78E-06 | -0.027 | 0.670 | Yes | AC069243.1;<br>EGFEM1P;<br>RP11-637O11.2 |
| 3:167935125 | rs6775766 | 0.343 | A/T | -0.328 | 3.26E-06 | -0.003 | 0.938 | Yes | NA |
| 17:3856615 | rs1800915 | 0.549 | A/G | 0.302 | 4.06E-06 | 0.024 | 0.446 | No | NA |
| 9:92497871 | rs28721393 | 0.218 | G/A | -0.358 | 6.90E-06 | -0.003 | 0.940 | Yes | RP5-1050E16.2;<br>RP5-1050E16.1 |
| 11:9578761 | rs72854168 | 0.181 | A/G | -0.406 | 7.47E-06 | -0.024 | 0.571 | Yes | NA |
| 9:132261992 | rs62584056 | 0.252 | A/G | -0.343 | 8.23E-06 | 0.019 | 0.591 | Yes | LINC00963;<br>RP11-492E3.51 |
| 9:99911969 | rs139074941 | 0.059 | A/G | 0.573 | 8.56E-06 | 0.028 | 0.663 | No | NA |
| 14:57195460 | rs140513548 | 0.050 | A/T | 0.625 | 8.99E-06 | 0.000 | 0.995 | Yes | RP11-1085N6.3 |

Marker: Chromosome + position based on NCBI Build 37 (hg19); A1/A2: effect allele and non-effect allele; Effect: beta-coefficient; S.E.: standard error; p-value GWAS: p-value of the same SNP in the genome-wide association study.

**Figure 2.**
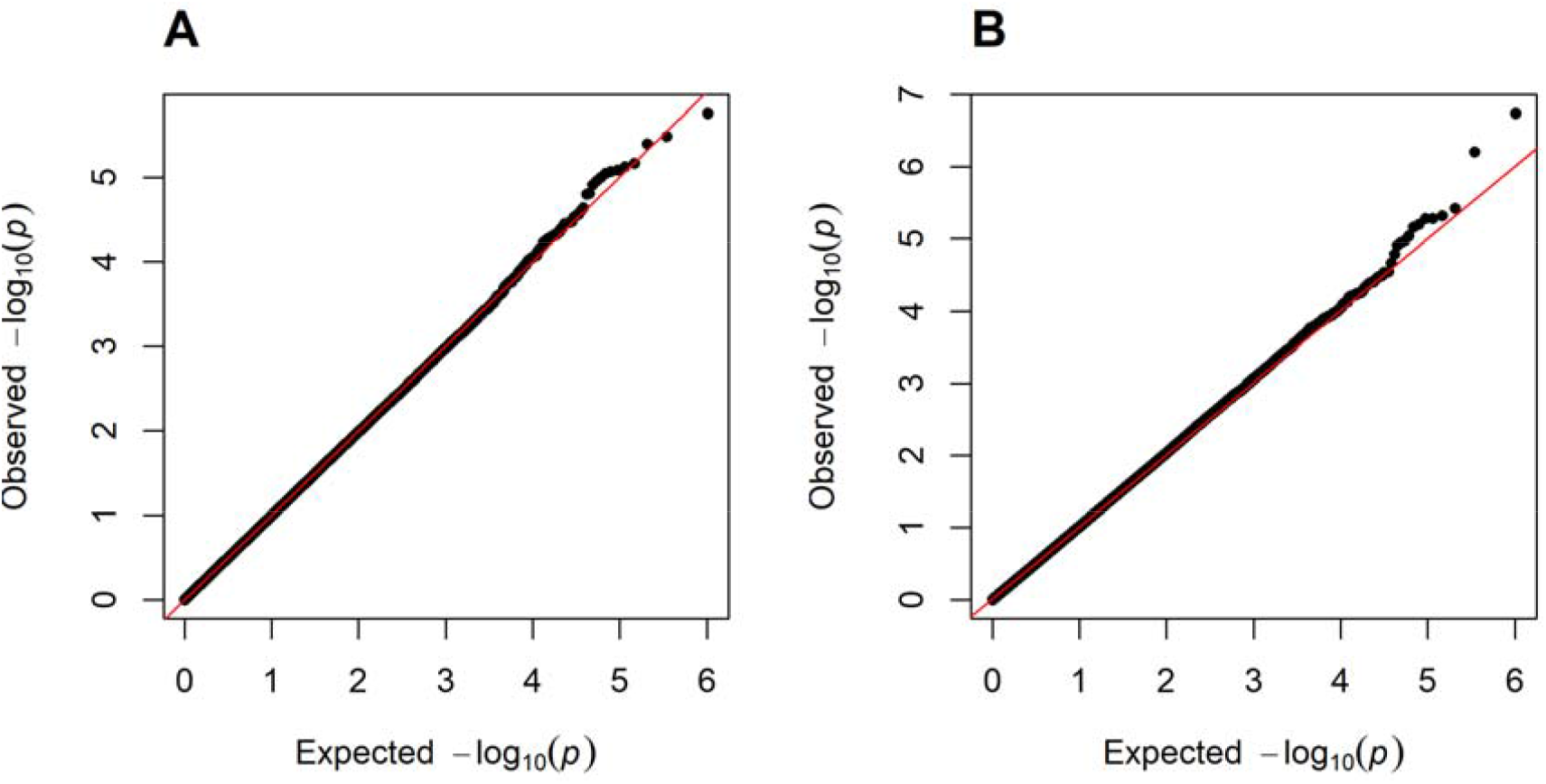
A and B: Quantile–quantile plot of genome-wide interaction (A) and genome-wide association (B) p-values.

### Genome-wide-association study findings

None of the SNPs from the GWAS reached genome-wide significance (Figure 1B). Table 3 summarizes the nine SNPs with a p-value < 1e-5 that were carried forward for Functional Mapping and Annotation analyses. Seven SNPs were independent lead SNPs and five of them were mapped to a gene. Figure 2B shows the QQ plots of observed versus expected p-values, which closely followed the null distribution (λ=1.02). The autosomal SNP-based heritability (h^2^_SNP_) on the liability scale was 0.06 (SE = 0.09).

**Table 3:**
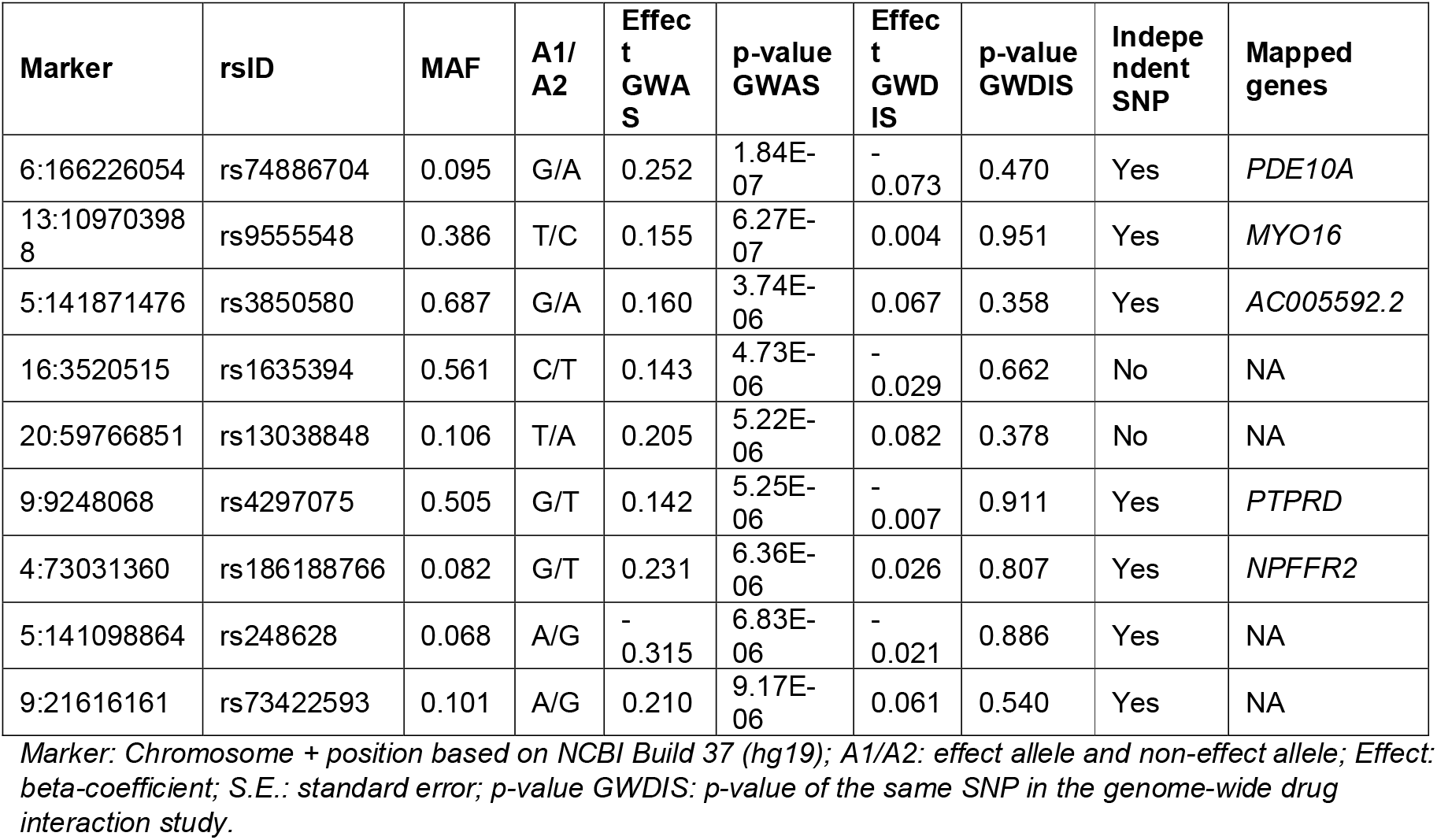
SNPS with a p-value < 1e-5 in the genome-wide association study on depression in young adulthood. None of the SNPs passed the genome-wide significance threshold (< 5e-8).

### Gene-level analysis

A gene-level analysis was performed using Multi-marker Analysis of GenoMic Annotation (MAGMA), which annotated 14798 genes (Supplementary Tables S3 and S4). None of the genes passed the genome-wide significance threshold, but some genes with a p-value < 1e-4 may be of potential interest. For the GWDIS, these were *FSIP1* (p = 5.90□×□10^−^□) and *EHBP1* (p = 6.47□×□10^−^□). For the GWAS, these were *MTMMR8* (p□=□ 8.59□×□10^−^□) and ZFYVE1 (p□=□9.57□×□10^−^□).

## Discussion

To better understand why some individuals may experience mood-based side effects of OC while others do not, we conducted a genome-wide study to identify genetic variants associated with depression and genetic variants that modify the association between OC and depression in young adulthood. No genetic variants or genes reached genome-wide significance in both the GWAS and the GWDIS.

The gene-level analyses of the GWDIS revealed two suggestive genes which both have not been associated with depression in previous studies.^57^ *FSIP1* has been found to be associated with gamma-glutamyl transferase,^58^ an enzyme with high intracellular concentrations in the liver which is increasingly viewed as a systemic marker of oxidative stress.^59,60^ Interestingly, research has shown that OC intake is associated with higher levels of γ-glutamyl transferase, suggesting that OC use leads to higher oxidative stress,^60–62^ which in turn may be implicated in neurodegenerative and neuropsychiatric disorders.^63^ The other suggestive gene, *EHBP1*, regulates vesicular trafficking and has been associated with body mass index, weight, coronary artery disease and low-density lipoprotein cholesterol.^64–67^

A prior GWAS showed that depression has a highly polygenic architecture involving many genetic variants with relatively small effects.^68^ The current largest GWAS found a polygenic score that explained only 5.8% of the variance.^5^ It is very likely that gene-OC interactions in the development of depression are polygenic in nature, just like depression itself.^68^ With a larger sample, it is possible that a polygenic score capturing many SNP-OC interactions on depression could be identified. However, our SNP-heritability based genome-wide interaction estimates suggest that even if found, the contribution to depression incidence is small, limiting its clinical applicability.

Some limitations should be taking into account when interpreting the results. First, OC use was based on retrospective self-report during the initial assessment visit, which is prone to recall bias. Second, N=165 participants reported OC initiation and depression diagnosis in the same year, for whom the order of occurrence could not be determined. We excluded these individuals from analysis, which may have influenced the findings. Third, being diagnosed with depression in adolescence or young adulthood was less common when most UK Biobank participants were young, so depression may have been underdiagnosed.^69^ Even with a relatively large total sample size of the study, only N=2,067 depression cases before the age of 25 years is considered small for a whole-genome interaction study. While low power may therefore explain the null findings in this study, the SNP-heritability based genome-wide interaction estimates generally favor interpreting the results as true null effect. Fourth, we did not have information on the type of OC used. Since different types of synthetic estrogens and progestins could potentially interact with different genetic factors, this may have obscured potential associations. However, the majority of the participants likely initiated the second-generation pills, with ethinyl-estradiol and levonorgestrel or norgestrel, which were predominantly used in the United Kingdom in 1970s and early 1980s.^34^

In conclusion, in this large population-based genome-wide study, we did not find evidence for genetic variants that modify the association between OC initiation and depression in young adulthood. If such effects exist, they are likely to be small and polygenic, or not well captured by current approaches. At present, there is insufficient evidence to support the use of genetic information for individualised contraception counselling with respect to mood-based side effects.

## Acknowledgements

This research has been conducted using the UK Biobank Resource under application number 80668. The analyses were conducted on the Research Analysis Platform (http://ukbiobank.dnanexus.com). We acknowledge the participants and staff involved in UKB for their contribution. During the preparation of this work the authors used generative AI to assist with debugging analysis scripts and improving sentence clarity. The authors reviewed and edited the content as needed and take full responsibility for the content of the published article.

## Author contributions

CAE, FC and RHM conceptualized and designed the study. CAE drafted the manuscript. All authors contributed to data interpretation and substantially revised the work.

## Data availability statement

The genetic and phenotype datasets generated by UK Biobank analysed during the current study are available via the UK Biobank data access process (see http://www.ukbiobank.ac.uk/register-apply/). Scripts and syntaxes used in this study can be found at https://github.com/centhoven/genome-wide-by-OC-interaction.

## References

1. Depression and Other Common Mental Disorders: Global Health Estimates. Geneva: World Health Organization vol. 24 (2017).

2. Moreno-Agostino, D. et al. Global trends in the prevalence and incidence of depression:a systematic review and meta-analysis. J. Affect. Disord. 281, 235–243 (2021).

3. Marx, W. et al. Major depressive disorder. Nat. Rev. Dis. Primers 9, 44 (2023).

4. Kuehner, C. Why is depression more common among women than among men? Lancet Psychiatry 4, 146–158 (2017).

5. Adams, M. J. et al. Trans-ancestry genome-wide study of depression identifies 697 associations implicating cell types and pharmacotherapies. Cell 188, 640–652.e9 (2025).

6. Silveira, P. P., Pokhvisneva, I., Howard, D. M. & Meaney, M. J. A sex-specific genome-wide association study of depression phenotypes in UK Biobank. Mol. Psychiatry 28, 2469–2479 (2023).

7. Rubinow, D. R. & Schmidt, P. J. Sex differences and the neurobiology of affective disorders. Neuropsychopharmacology 44, 111–128 (2019).

8. Contraceptive Use by Method 2019. (UN, 2019). doi:10.18356/1bd58a10-en.

9. Porcu, P., Serra, M. & Concas, A. The brain as a target of hormonal contraceptives: Evidence from animal studies. Front. Neuroendocrinol. 55, 100799 (2019).

10. Skovlund, C. W., Mørch, L. S., Kessing, L. V., Lange, T. & Lidegaard, Ø. Association of Hormonal Contraception With Suicide Attempts and Suicides. American Journal of Psychiatry 175, 336–342 (2018).

11. Anderl, C., Li, G. & Chen, F. S. Oral contraceptive use in adolescence predicts lasting vulnerability to depression in adulthood. Journal of Child Psychology and Psychiatry 61, 148–156 (2020).

12. Anderl, C., de Wit, A. E., Giltay, E. J., Oldehinkel, A. J. & Chen, F. S. Association between adolescent oral contraceptive use and future major depressive disorder: a prospective cohort study. Journal of Child Psychology and Psychiatry 63, 333–341 (2022).

13. Skovlund, C. W., Mørch, L. S., Kessing, L. V. & Lidegaard, Ø. Association of Hormonal Contraception With Depression. JAMA Psychiatry 73, 1154 (2016).

14. de Wit, A. E. et al. Hormonal contraceptive use and depressive symptoms: systematic review and network meta-analysis of randomised trials. BJPsych Open 7, e110 (2021).

15. Duke, J. M., Sibbritt, D. W. & Young, A. F. Is there an association between the use of oral contraception and depressive symptoms in young Australian women? Contraception 75, 27–31 (2007).

16. McKetta, S. & Keyes, K. M. Oral contraceptive use and depression among adolescents. Ann. Epidemiol. 29, 46–51 (2019).

17. Doornweerd, A. M. et al. Stable Anxiety and Depression Trajectories in Late Adolescence for Oral Contraceptive Users. Front. Psychiatry 13, (2022).

18. Keyes, K. M. et al. Association of hormonal contraceptive use with reduced levels of depressive symptoms: A national study of sexually active women in the United States. Am. J. Epidemiol. 178, 1378–1388 (2013).

19. Bosmans, N. H. M. et al. Associations between oral hormonal contraceptives and internalising problems in adolescent girls. BJPsych Open 11, e40 (2025).

20. Chen, F. S., Zareian, B., Nelson, M. A., Edwards, N. & Anderl, C. Research Review: Are sampling biases masking long-term effects of hormonal contraceptive use in adolescence on risk for depression? Journal of Child Psychology and Psychiatry https://doi.org/10.1111/jcpp.14180 (2025) doi:10.1111/jcpp.14180.

21. Larsen, S. V., Frokjaer, V. G. & Ozenne, B. Methodological obstacles in studies linking hormonal contraception and depression. The British Journal of Psychiatry 226, 334–336 (2025).

22. Hill, S. E. & Mengelkoch, S. Moving beyond the mean: Promising research pathways to support a precision medicine approach to hormonal contraception. Front. Neuroendocrinol. 68, 101042 (2023).

23. Pérez-López, F. R., Pérez-Roncero, G. R., López-Baena, M. T., Santabárbara, J. & Chedraui, P. Hormonal contraceptives and the risk of suicide: a systematic review and meta-analysis. European Journal of Obstetrics & Gynecology and Reproductive Biology 251, 28–35 (2020).

24. Davies-Kellock, M., Potts, H. W., Pinho, L. G. B., Chilanga, F. & Farič, N. Association of Oral Contraceptives with Depression Symptoms, Diagnosis, and Treatment in Healthy Women: A Meta-Analysis. Preprint at 10.2139/ssrn.6019235 (2026).

25. Kraft, M. Z. et al. Symptoms of mental disorders and oral contraception use: A systematic review and meta-analysis. Front. Neuroendocrinol. 72, 101111 (2024).

26. Kellock, M. D., Potts, H. W. W., Pinho, G., Chilanga, F. & Farič, N. Association of oral contraceptives with depression symptoms, diagnosis, and treatment in healthy women: A meta-analysis. European Journal of Obstetrics & Gynecology and Reproductive Biology 323, 115144 (2026).

27. Goldzieher, J. W. & Stanczyk, F. Z. Oral contraceptives and individual variability of circulating levels of ethinyl estradiol and progestins. Contraception 78, 4–9 (2008).

28. Lin, Y., Anderson, G. D., Kantor, E., Ojemann, L. M. & Wilensky, A. J. Differences in the urinary excretion of 6-β-hydroxycortisol/cortisol between Asian and Caucasian women. The Journal of Clinical Pharmacology 39, 578–582 (1999).

29. Klok, M. D. et al. A common and functional mineralocorticoid receptor haplotype enhances optimism and protects against depression in females. Transl. Psychiatry 1, e62–e62 (2011).

30. Hamstra, D. A., de Kloet, E. R., van Hemert, A. M., de Rijk, R. H. & Van der Does, A. J. W. Mineralocorticoid receptor haplotype, oral contraceptives and emotional information processing. Neuroscience 286, 412–422 (2015).

31. Winham, S. J. & Biernacka, J. M. Gene–environment interactions in genome-wide association studies: current approaches and new directions. Journal of Child Psychology and Psychiatry 54, 1120–1134 (2013).

32. Bycroft, C. et al. The UK Biobank resource with deep phenotyping and genomic data. Nature 562, 203–209 (2018).

33. Hanscombe, K. B., Coleman, J. R. I., Traylor, M. & Lewis, C. M. ukbtools: An R package to manage and query UK Biobank data. PLoS One 14, e0214311 (2019).

34. Johansson, T. et al. Population-based cohort study of oral contraceptive use and risk of depression. Epidemiol. Psychiatr. Sci. 32, e39 (2023).

35. First Occurrence of Health Outcomes Defined by 3-Character ICD10 Code. UK Biobank, Stockport (2019).

36. Keyes, K. M. et al. Age, Period, and Cohort Effects in Psychological Distress in the United States and Canada. Am. J. Epidemiol. 179, 1216–1227 (2014).

37. Freeman, A. et al. The role of socio-economic status in depression: results from the COURAGE (aging survey in Europe). BMC Public Health 16, 1098 (2016).

38. Hirtz, R. et al. Causal Effect of Age at Menarche on the Risk for Depression: Results From a Two-Sample Multivariable Mendelian Randomization Study. Front. Genet. 13, (2022).

39. Jiang, L. et al. Is early menarche related to depression? A meta-analysis. J. Affect. Disord. 369, 508–515 (2025).

40. Slaymaker, E. et al. Trends in sexual activity and demand for and use of modern contraceptive methods in 74 countries: a retrospective analysis of nationally representative surveys. Lancet Glob. Health 8, e567–e579 (2020).

41. Xi, D., Chen, B., Tao, H., Xu, Y. & Chen, G. The risk of depressive and anxiety symptoms in women with premature ovarian insufficiency: a systematic review and meta-analysis. Arch. Womens Ment. Health 26, 1–10 (2023).

42. Zehravi, M., Maqbool, M. & Ara, I. Depression and anxiety in women with polycystic ovarian syndrome: a literature survey. Int. J. Adolesc. Med. Health 33, 367–373 (2021).

43. Moore, J., Kennedy, S. & Prentice, A. Modern combined oral contraceptives for pain associated with endometriosis. in Cochrane Database of Systematic Reviews (ed. Kennedy, S.) (John Wiley & Sons, Ltd, Chichester, UK, 1997). doi:10.1002/14651858.CD001019.

44. Gambadauro, P., Carli, V. & Hadlaczky, G. Depressive symptoms among women with endometriosis: a systematic review and meta-analysis. Am. J. Obstet. Gynecol. 220, 230–241 (2019).

45. Zhao, S., Wu, W., Kang, R. & Wang, X. Significant Increase in Depression in Women With Primary Dysmenorrhea: A Systematic Review and Cumulative Analysis. Front. Psychiatry 12, (2021).

46. Davis, A. R. & Westhoff, C. L. Primary Dysmenorrhea in Adolescent Girls and Treatment with Oral Contraceptives. J. Pediatr. Adolesc. Gynecol. 14, 3–8 (2001).

47. Chang, C. C. et al. Second-generation PLINK: rising to the challenge of larger and richer datasets. Gigascience 4, (2015).

48. Carss, K. et al. Whole-genome sequencing of 490,640 UK Biobank participants. Nature 645, 692–701 (2025).

49. Josse, J. & Husson, F. missMDA: A Package for Handling Missing Values in Multivariate Data Analysis. J. Stat. Softw. 70, 1–31 (2016).

50. Therneau, T. M. & Grambsch, P. M. Modeling Survival Data: Extending the Cox Model. (Springer New York, New York, NY, 2000). doi:10.1007/978-1-4757-3294-8.

51. Watanabe, K., Taskesen, E., van Bochoven, A. & Posthuma, D. Functional mapping and annotation of genetic associations with FUMA. Nat. Commun. 8, 1826 (2017).

52. Lawrence, M., Gentleman, R. & Carey, V. rtracklayer: an R package for interfacing with genome browsers. Bioinformatics 25, 1841–2 (2009).

53. Perez, G. et al. The UCSC Genome Browser database: 2025 update. Nucleic Acids Res. 53, D1243–D1249 (2025).

54. de Leeuw, C. A., Mooij, J. M., Heskes, T. & Posthuma, D. MAGMA: generalized gene-set analysis of GWAS data. PLoS Comput. Biol. 11, e1004219 (2015).

55. Kanai, M. et al. Genetic analysis of quantitative traits in the Japanese population links cell types to complex human diseases. Nat. Genet. 50, 390–400 (2018).

56. Brennan, P. N., Dillon, J. F. & Tapper, E. B. Gamma-Glutamyl Transferase (γ-GT) – an old dog with new tricks? Liver International 42, 9–15 (2022).

57. Kowalska, K. & Milnerowicz, H. Pro/antioxidant status in young healthy women using oral contraceptives. Environ. Toxicol. Pharmacol. 43, 1–6 (2016).

58. Nilssen, O., Førde, O. H. & Brenn, T. The Tromsø Study. Distribution and population determinants of gamma-glutamyltransferase. Am. J. Epidemiol. 132, 318–26 (1990).

59. Straus, B., Calic-runje, R. & Cepelak, I. Influence of contraceptives on gamma glutamyltransferase activity. Acta Pharm. Jugosl. 32, 191–5 (1982).

60. Salim, S. Oxidative Stress and the Central Nervous System. J. Pharmacol. Exp. Ther. 360, 201–205 (2017).

61. Rai, A., Bleimling, N., Vetter, I. R. & Goody, R. S. The mechanism of activation of the actin binding protein EHBP1 by Rab8 family members. Nat. Commun. 11, 4187 (2020).

62. Siewert, K. M. & Voight, B. F. Bivariate Genome-Wide Association Scan Identifies 6 Novel Loci Associated With Lipid Levels and Coronary Artery Disease. Circ. Genom. Precis. Med. 11, e002239 (2018).

63. Pulit, S. L. et al. Meta-analysis of genome-wide association studies for body fat distribution in 694 649 individuals of European ancestry. Hum. Mol. Genet. 28, 166–174 (2019).

64. Watanabe, K. et al. A global overview of pleiotropy and genetic architecture in complex traits. Nat. Genet. 51, 1339–1348 (2019).

65. McIntosh, A. M., Sullivan, P. F. & Lewis, C. M. Uncovering the Genetic Architecture of Major Depression. Neuron 102, 91–103 (2019).

66. Klerman, G. L. The Current Age of Youthful Melancholia. British Journal of Psychiatry 152, 4–14 (1988).

