## Supplemental Tables 1-2 for "Interactive effects of genetic variants and oral contraceptive use on depression in the UK Biobank"

**Supplementary material**

**Supplementary Table 1: Deviations from the pre-registration**

| **Pre-registration** | **Deviations** |
| --- | --- |
| *We will perform a cox regression model with OC use on incident depression.* | The proportional hazard assumption was violated when using a cox proportional hazards model with OC ever versus never use. Therefore, we used a cox proportional hazards model for count data with time dependent covariates to account for age at OC initiation, age at menarche, age at sexual debut and age at first acne diagnosis.^50^ |
| *The participants will be followed from birth until the first occurrence of depression, if they reached menopause (UKB field: 3581), underwent a hysterectomy (UKB field: 2824) or a bilateral oophorectomy (UKB field: 3882), or the end of follow-up (age at UKB assessment visit; UKB field: 21022) whichever came first. A previous study using UK Biobank data showed that the effect of OC use on depression is highest after two years of OC use, therefore OC users will be censored two years after OC initiation.^3^* | We followed participants from birth until the first occurrence of depression, when they became pregnant (age at first birth minus 1), if they reached menopause, underwent a hysterectomy, a bilateral oophorectomy or when they turned 25 years old, whichever came first in order to harmonize the data of the discovery sample (UK Biobank) with the data of the adolescents from the validation sample (Generation R). This approach allowed us to examine OC initiation in adolescence and young adulthood, and therefore we did not censor participants two years after OC initiation. |
| *All analyses will be adjusted for year of birth (UKB field: 34),^8^ Townsend deprivation index (proxy for socioeconomic status; UKB field: 22189),^9,10^ age at menarche (UKB field: 2714),^17,18^ age at sexual debut (UKB field: 2139),^10,19^ ovarian dysfunction (UKB field: 130736),^11,12^ endometriosis (UKB field: 132122),^13,14^ dysmenorrhea (UKB field: 132146),^15,16^ the first eight genetic principal components (UKB field: 22009),^2^ and genotyping array (for the GWAS and GWDIS; UKB field:* *22000).* | We additionally adjusted for age at first reported acne (UKB field: 131790). |
| *For the GWAS and the GWDIS, the software REGENIE will be used to conduct cox regression for time to event data.^7^* | We could not use REGENIE because it does not allow time dependent covariates. We therefore conducted all analyses in R. |
| *Variants with a minor allele frequency <1%, or an imputation accuracy Info score <0.8 will be removed.^6^* | Variants with a minor allele frequency <5% were removed to reduce the possibility of false positive tests. We additionally used pruning to reduce the total number of SNPs to be tested to reduce computational power and time. |

**Supplementary Table 2: Descriptions about the assessment of the covariates in the UK Biobank.**

| **Variables** | **Field numbers** | **Description** |
| --- | --- | --- |
| Year of birth | p34 | Year of birth was acquired from central registry. |
| Townsend deprivation index | p22189 | The Townsend deprivation index was used as a proxy for socioeconomic status. It was based on the preceding national census output areas, corresponding to each participants’ postcode. |
| Age at menarche | p2714_i0  p2714_i1  p2714_i2  p2714_i3 | Based on a touchscreen question "How old were you when your periods started?". The answer options “Do not know” and “Prefer not to answer” were set to missing. The question was asked during the initial assessment visit (i0), first repeat assessment visit (i1), imaging visit (i2) and/or first repeat imaging visit (i3). In case the data from the initial assessment visit was missing, the data from the first repeat assessment visit was used, and so forth. |
| Age at sexual debut | p2139_i0  p2139_i1  p2139_i2  p2139_i3 | Based on a touchscreen question "What was your age when you first had sexual intercourse? (Sexual intercourse includes vaginal, oral or anal intercourse)". The answer options “Do not know” and “Prefer not to answer” were set to missing. The answer option “Never had sex” (0.9% of the participants) was set to the age at assessment in order to process this variable as continuous. The question was asked during the initial assessment visit (i0), first repeat assessment visit (i1), imaging visit (i2) and/or first repeat imaging visit (i3). In case the data from the initial assessment visit was missing, the data from the first repeat assessment visit was used, and so forth. |
| Age at first reported acne | p131790 | Date of the first occurrence of any code mapped to 3-character ICD10 L70 (acne). Age at first reported acne was calculated by subtracting the participants year of birth (UKB field: p34) from the year of first reported acne. |
| Ovarian dysfunction | p130736 | Date of the first occurrence of any code mapped to 3-character ICD10 E28 (ovarian dysfunction). |
| Endometriosis | p132122 | Date of the first occurrence of any code mapped to 3-character ICD10 N80 (endometriosis). Categorized into ‘yes’ versus ‘no’. |
| Dysmenorrhea | p132146 | Date of the first occurrence of any code mapped to 3-character ICD10 N92 (excessive, frequent and irregular menstruation). Categorized into ‘yes’ versus ‘no’. |
| Genetic principal components | p22009_a1  p22009_a2  p22009_a3  p22009_a4  p22009_a5  p22009_a6  p22009_a7  p22009_a8 | Principal components were calculated using the fastPCA algorithm using a sample of N=407,219 unrelated, high quality samples and 147,604 high quality markers. Following, the principal component-loadings were computed for all samples in the cohort. |
| Genotype measurement batch | p22000 | Batch was categorized into the UK BiLEVE Axiom array and the UK Biobank Axiom array. |

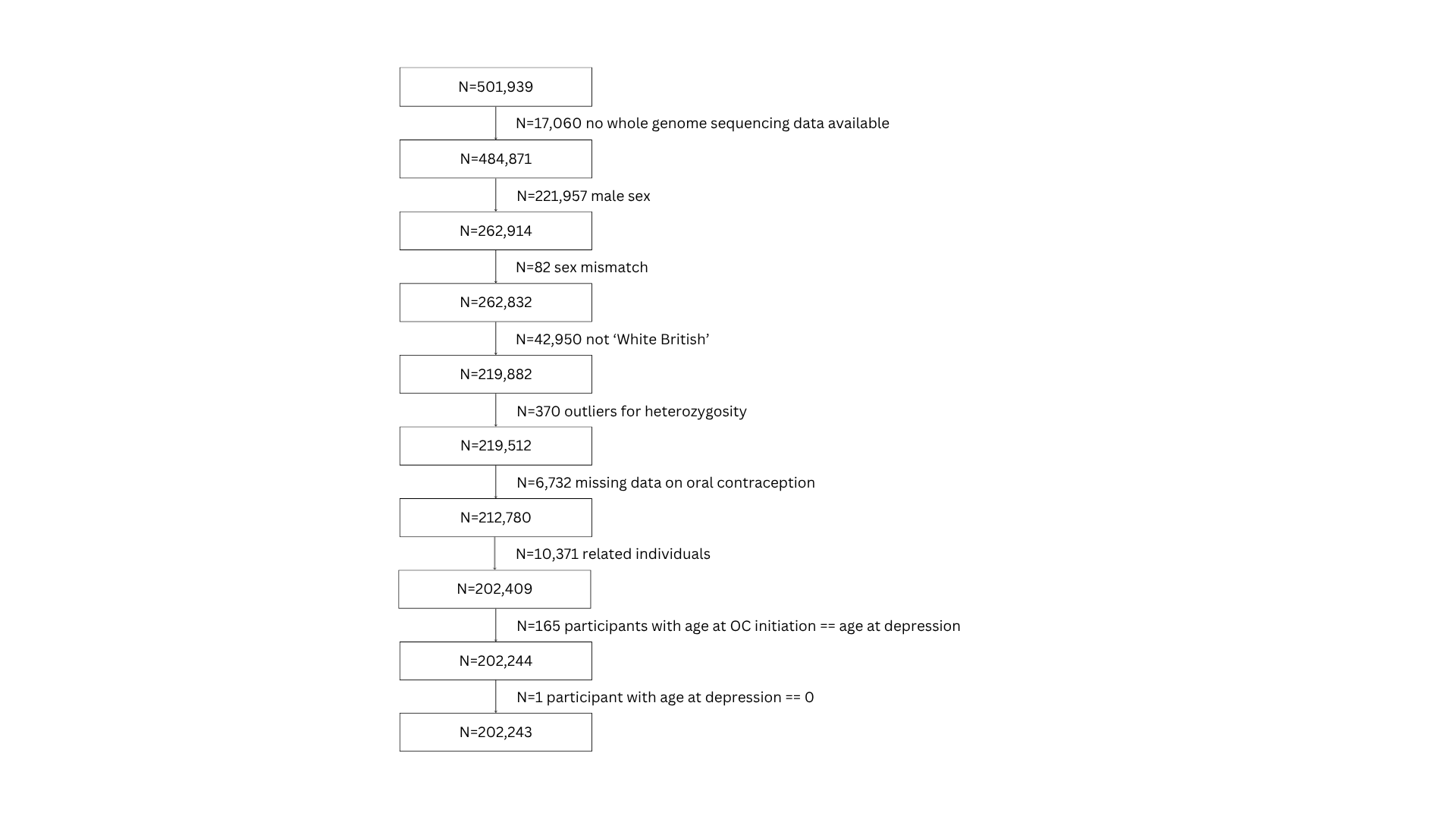

**Supplementary Figure 1: Flowchart of participants from the UK Biobank study population included in analyses**
